# Bridging Policy and Practice: Parents’ and Caregivers’ Experiences with the Interim Canada Dental Benefit in Canada

**DOI:** 10.64898/2026.05.12.26352368

**Authors:** Olubukola O. Olatosi, Thiago H.L. Baltus, Betty-Anne Mittermuller, Silvana Fux, Athena Monayao, JuHae Lee, Anil Menon, Katherine Yerex, Saif Goubran, Robert J. Schroth

## Abstract

**Background:** Access to dental care remains a significant challenge for many children in Canada, particularly among low-income and underserved populations. The Interim Canada Dental Benefit (CDB), introduced in October 2022, aimed to reduce financial barriers to oral health care for children under 12 years of age while the Canadian Dental Care Plan (CDCP) was being developed. While emerging evidence has examined program uptake, limited qualitative research has explored parents’ and caregivers’ experiences with the Interim CDB.

**Objective:** This study aimed to explore parents’ and caregivers’ experiences with the Interim CDB in Manitoba, Canada, including awareness, access, perceived benefits, challenges, and recommendations for program improvement.

**Methods:** A qualitative descriptive study was conducted using semi-structured interviews with 30 parents and caregivers of children under 12 years of age. Participants were recruited primarily through community dental clinics. Interviews were conducted between July 2023 and February 2024, audio-recorded, and transcribed verbatim. Data were analyzed using inductive thematic analysis to identify key themes and subthemes.

**Results:** Seven interconnected themes were identified: (1) limited and uneven awareness of the Interim CDB; (2) inadequate and inequitable communication strategies; (3) barriers to accessing the benefit, including misconceptions about eligibility and complex application processes; (4) dental providers as key facilitators of access; (5) financial relief and improved access to care; (6) gaps in coverage and ongoing financial strain; and (7) participant-driven recommendations for improvement. While the benefit was widely perceived as reducing financial barriers and enabling access to care, challenges related to awareness, communication, and adequacy of coverage limited its overall effectiveness. Participants emphasized the need for improved communication from government, simplified application processes, expanded eligibility, and increased financial support.

**Conclusion:** The Interim CDB represents an important step toward improving access to dental care for children in Canada. However, this study highlights critical implementation gaps related to awareness, accessibility, and coverage. Addressing these challenges will be essential to ensuring the success of the new CDCP and advancing equitable access to oral health care.

## Introduction

Oral health is a fundamental component of overall health and well-being, yet access to dental care remains a persistent challenge for many children and families (Han et al., 2025; Menon et al., 2026). Dental caries continues to be one of the most prevalent chronic conditions affecting children globally, disproportionately impacting those from low-income and underserved populations (Tungare & Paranjpe, 2025). Financial barriers, lack of dental insurance, and limited access to publicly funded dental services contribute significantly to unmet oral health needs and delayed care (Gondro et al., 2025).

In Canada, dental care is largely delivered through a privately financed system, which creates inequities in access for families without employer-sponsored insurance or sufficient financial resources (Menon et al., 2024). Although publicly funded programs exist, they are often limited in scope, fragmented across jurisdictions, and insufficient to meet population needs. As a result, many families incur out-of-pocket expenses, which can delay or prevent timely dental care for children (Levy et al., 2023).

In response to these longstanding gaps, the Government of Canada introduced the Interim Canada Dental Benefit (CDB) in October 2022. This temporary program was designed to provide direct financial support to eligible low-income families with children under 12 years of age, while the Canadian Dental Care Plan (CDCP) is implemented, with the broader goal of improving access to dental care. The Interim CDB represented a significant federal policy shift toward addressing oral health inequities and expanding access to care.

The Interim CDB targeted children under 12 years of age from families with annual income of less than $90,000 who did not have access to private dental insurance (Baltus et al., 2026a; Goubran et al., 2026). Eligibility required families to be Canadian citizens or permanent residents of Canada, to have filed a recent tax return with the Canada Revenue Agency (CRA), and to have incurred out-of-pocket dental expenses within the program period(Canada Revenue Agency, 2022). The benefit was distributed in two periods: Period 1, from October 2022 to June 30, 2023, and Period 2, from July 1, 2023, to June 30, 2024. Families with private dental insurance could not apply for the benefit, however those holding some governmental dental insurance (e.g., Employment and Income Assistance; Non-Insured Health Benefits) were still eligible. Financial support ranged from $260 to $650 per child, depending on adjusted family income, as outlined under Bill C-31 (Government of Canada. Government Bill (House of Commons), 2022). The program operated as a precursor to the CDCP and sunsetted on June 30, 2024, with the CDCP subsequently launched to provide broader coverage for uninsured Canadians.

Emerging evidence has begun to examine the early implementation and impact of the Interim CDB (Goubran et al., 2024; Goubran et al., 2026; Menon et al., 2026; Schroth et al., 2023; Schroth et al., 2025). Analyses of program uptake suggest that while the benefit has reached a substantial number of eligible children, disparities in utilization persist, particularly among uninsured and socioeconomically disadvantaged populations (Goubran et al., 2024; Goubran et al., 2026; Schroth et al., 2023). Quantitative investigations of program data have identified trends in uptake during the first year, the first 18 months, and the full 21 months of implementation, including lower participation rates in certain regions such as the Territories. Complementary research exploring oral health professionals’ perspectives has identified both opportunities and challenges in program delivery, including concerns about awareness, administrative processes, and integration within existing care systems (Baltus et al., 2026a).

Recent work examining parental perspectives has also underscored ongoing barriers to accessing dental care, including cost, limited service availability, and challenges in navigating benefit programs (Menon et al., 2026). However, despite these important contributions, there remains limited in-depth qualitative evidence exploring how parents and caregivers experience the Interim CDB in practice, particularly with respect to awareness, communication, access, and perceived impact on children’s oral health care.

Understanding these lived experiences is critical, as the effectiveness of publicly funded dental programs depends not only on financial coverage but also on equitable access, clear communication, and user-friendly implementation. Qualitative approaches are uniquely positioned to capture nuanced perspectives and identify system-level gaps that may not be apparent in quantitative analyses alone.

Therefore, this study aimed to explore parents’ and caregivers’ experiences with the Interim CDB in Manitoba, Canada. Specifically, this study sought to examine awareness and understanding of the program, perceptions of communication and information delivery, barriers and facilitators to accessing the benefit, perceived impact on children’s oral health care, and participant-driven recommendations to inform the ongoing implementation of the CDCP.

## Methods

### Study Design

We conducted a qualitative descriptive study to explore parents’ and caregivers’ experiences with the Interim CDB. A qualitative descriptive approach was selected as it allows for a comprehensive, low-inference summary of participant perspectives and is well-suited for informing health policy and program implementation.

### Participants and Recruitment

Participants were parents or caregivers (referred to as “parents” for the remainder of this paper) of children under 12 years of age who had experience with the Interim CDB, including those who had heard of, applied for, used, or chosen not to use the benefit. A purposive sampling strategy was used to capture a range of experiences and perspectives. Participants were recruited primarily through community dental clinics, where study information was shared with eligible families. Additional recruitment occurred through word-of-mouth. Interested individuals contacted the research team and were screened for eligibility.

A total of 30 parents participated in the study. Sociodemographic information, including parental role, education, employment status, and insurance coverage, was collected to contextualize participant responses.

### Data Collection

Data were collected through semi-structured interviews conducted via telephone or virtual platforms, based on participant preference from July 2023 through February 2024. An interview guide was developed by the study team and stakeholders, in collaboration with Health Canada’s Oral Health Branch, to explore awareness, experiences, perceived benefits, challenges, and recommendations related to the Interim CDB. The interview guide consisted of 11 questions along with prompts (supplementary file) Interviews were conducted by a trained member of the research team with experience in qualitative research. Each interview lasted approximately 20– 45 minutes. All interviews were audio-recorded with participant consent and transcribed verbatim. Field notes were taken during and immediately after interviews to capture contextual observations and emerging insights.

### Data Analysis

Data were analyzed using thematic analysis. Transcripts were reviewed multiple times to achieve familiarity with the data. An initial coding framework was created inductively based on recurring patterns and concepts identified within the data.

Coding was conducted iteratively, with codes refined and grouped into broader categories through constant comparison. Related categories were then organized into overarching themes and subthemes that captured shared meanings across participants’ experiences.

To enhance analytical rigour, coding decisions and theme development were discussed within the research team. Discrepancies were resolved through discussion and consensus. Data analysis was supported by maintaining an audit trail of coding decisions and theme development.

### Ethics approval

Ethical approval was obtained from the University of Manitoba Health Research Ethics Board: HS26028 (H2023:180).

## Results

### Participant Characteristics

A total of 30 parents participated in the qualitative interviews. Most participants were female and identified as mothers, with the majority residing in urban settings. Participants represented diverse socio-demographic backgrounds, including Indigenous, African, Asian, and Caucasian populations. Most parents reported not having private dental insurance, highlighting the relevance of publicly funded dental support programs for this population (Table 1)

**Table 1.**
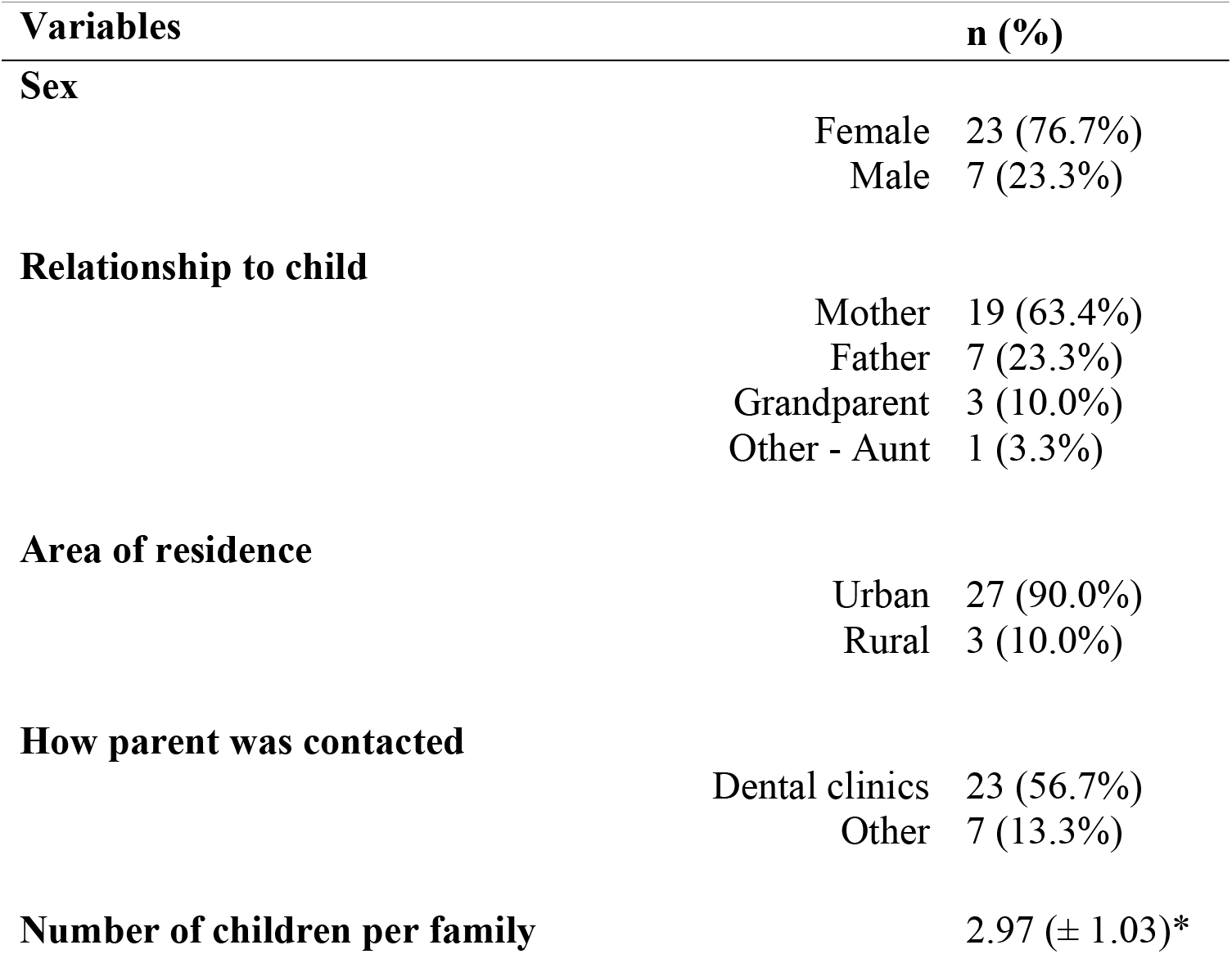
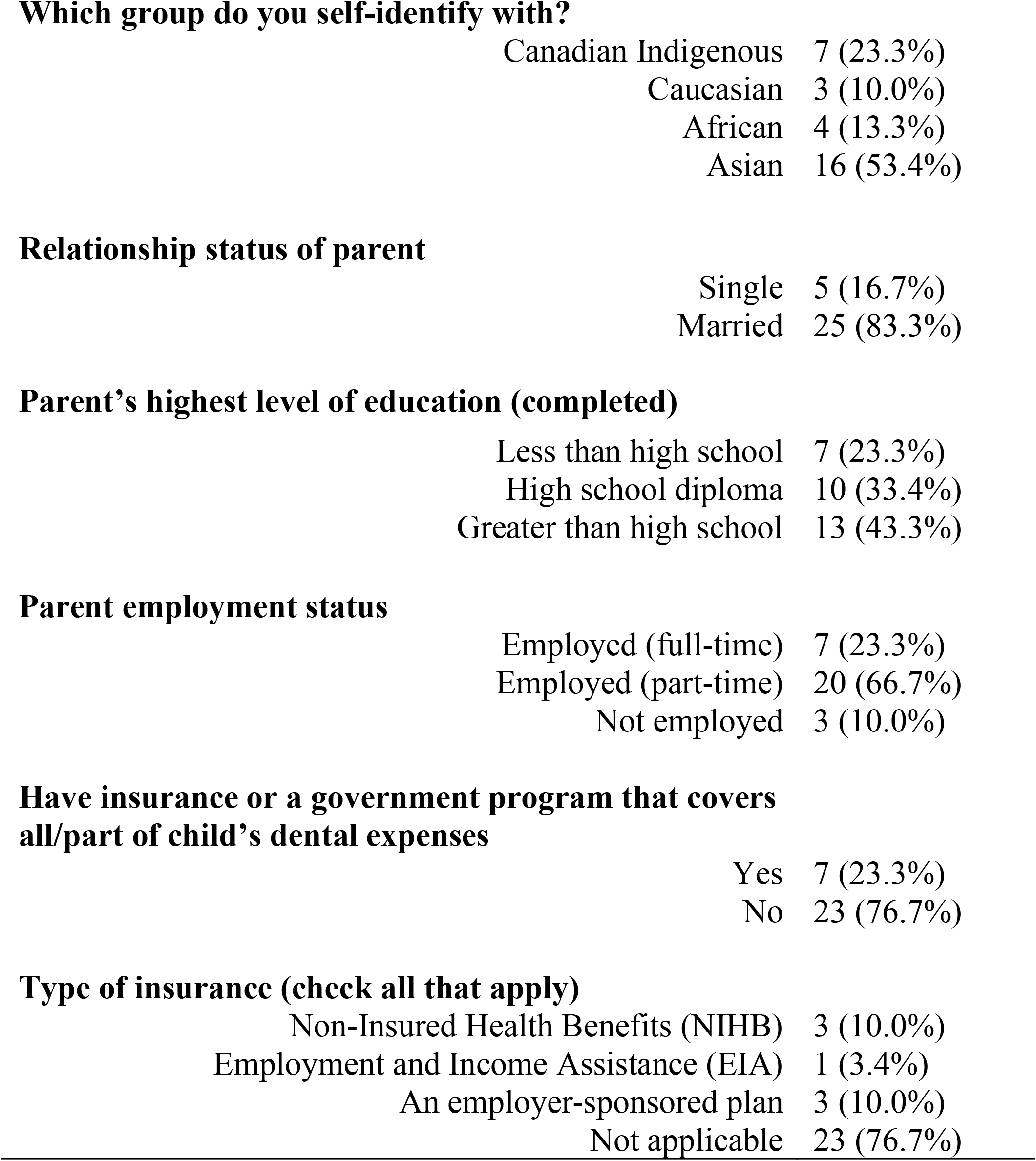
Parent Sociodemographic Results – Qualitative Interviews.

| **Variables** | **n (%)** |
| --- | --- |
| **Sex** |  |
| Female | 23 (76.7%) |
| Male | 7 (23.3%) |
| **Relationship to child** |  |
| Mother | 19 (63.4%) |
| Father | 7 (23.3%) |
| Grandparent | 3 (10.0%) |
| Other - Aunt | 1 (3.3%) |
| **Area of residence** |  |
| Urban | 27 (90.0%) |
| Rural | 3 (10.0%) |
| **How parent was contacted** |  |
| Dental clinics | 23 (56.7%) |
| Other | 7 (13.3%) |
| **Number of children per family** | 2.97 (± 1.03)* |
| **Which group do you self-identify with?** |  |
| Canadian Indigenous | 7 (23.3%) |
| Caucasian | 3 (10.0%) |
| African | 4 (13.3%) |
| Asian | 16 (53.4%) |
| **Relationship status of parent** |  |
| Single | 5 (16.7%) |
| Married | 25 (83.3%) |
| **Parent's highest level of education (completed)** |  |
| Less than high school | 7 (23.3%) |
| High school diploma | 10 (33.4%) |
| Greater than high school | 13 (43.3%) |
| **Parent employment status** |  |
| Employed (full-time) | 7 (23.3%) |
| Employed (part-time) | 20 (66.7%) |
| Not employed | 3 (10.0%) |
| **Have insurance or a government program that covers all/part of child's dental expenses** |  |
| Yes | 7 (23.3%) |
| No | 23 (76.7%) |
| **Type of insurance (check all that apply)** |  |
| Non-Insured Health Benefits (NIHB) | 3 (10.0%) |
| Employment and Income Assistance (EIA) | 1 (3.4%) |
| An employer-sponsored plan | 3 (10.0%) |
| Not applicable | 23 (76.7%) |

### Overview of themes

Analysis of participant responses generated seven interconnected themes:

1. Limited and uneven awareness of the Interim CDB
2. Inadequate and inequitable government communication strategies
3. Barriers to accessing the benefit
4. Dental providers as key facilitators
5. Financial relief and improved access to care
6. Gaps in coverage and ongoing financial strain
7. Participant-driven recommendations for improvement

Additional illustrative quotes supporting each theme and subtheme are presented in Table 2.

**Table 2.** Illustrative Quotes Supporting themes.

| **Themes** | **Subthemes** | **Illustrative Quotes** | **Participant ID** |
| --- | --- | --- | --- |
| **Limited and Uneven Awareness of the Interim CDB** | Variable levels of knowledge | “As far as I know, it's just... like the help... to pay for... if they're not insured.” | Parent #2SC |
|  | Partial understanding of purpose | “I know that it is a benefit... designed to help low-income families get to the dentist.” | Parent #3AR |
|  | Informal information sources | “Doctor B told me about it.” | Parent #2SC |
|  |  | “We visited the dental clinic... and at the reception we heard about the program.” | Parent #13GUD |
|  | inequities in awareness | “People that are poor definitely don't have Internet access... how would the people that need it most know about it?” | Participant #8CK |
| **Inadequate and Inequitable Communication Strategies** | Limited visibility | “I haven't heard anybody talk about it... I haven't seen any information on it.” | Parent #3AR |
|  | Poor communication approach | “They gave it out by email... not everybody has access to Internet.” | Participant #8CK |
|  | Confusing information | “CRA letter too complex and confusing.” | Multiple participants |
| **Barriers to Accessing the Interim CDB** | Misunderstood eligibility | “I wasn't eligible because... we're under the Treaty right now.” | Parent #7NT |
|  | Complex application process | "It is not something easy even for someone who is knowledgeable... what about the majority?" | Parent #4DG |
|  | System navigation challenges | "...last time my wife was trying to apply by phone and she was waiting, waiting, long time...I don't know if she completed it | Parent #19ST |
|  | Timing barriers | "I didn't know there was a time period... I had to pay out of my pocket." | Parent #15JB |
| **Dental Providers as Key Facilitators of Access** | Information support from clinics | "They informed me... I got a lot of information out of it." | Parent #5OM |
|  | Trust in providers | "Doctor Bob and the staff... 100% helpful." | Parent #9TT |
| **Financial Relief and Improved Access to Care** | Reduced financial burden | "It helped... took the burden out of my pocket." | Parent #15JB |
|  | Improved access | "It helps us a lot... we pay lesser... it helped our family." | Parent #26RR |
| **Gaps in Coverage and Ongoing Financial Strain** | Insufficient coverage | "It doesn't completely cover everything... just the cavity and cleaning." | Parent #15JB |
|  | Variability in adequacy | "For my daughter... it wasn't enough... for my son, it's okay." | Parent #4DG |
|  | Uncertainty about scope | "Is it per child or for all the kids?" | Parent #13GUD |
| **Participant-Driven Recommendations for Program Improvement** | Expand age eligibility | "Include children... up to eighteen... because they, they have a lot more problems than kids" | Parent #2SC<br>Parent #11KB |
|  |  | “Children over twelve... they need more things like braces.” |  |
| | Increase financial support | “They should increase the amount... maybe around \$1,000.” | Parent #14MB |
|  | Improve awareness | “It should be given out by mail... posted in all dental offices.” | Parent #8CK |
|  | Simplify and improve accessibility | “Application... not easy... especially for people with less education.” | Parent #4DG |

### Theme 1: Limited and Uneven Awareness of the Interim CDB

Participants demonstrated varying levels of awareness and understanding of the Interim CDB, ranging from no prior knowledge to partial or clear understanding. While some participants were unfamiliar with the program, most had a general sense that it was intended to support low-income families accessdental care. However, detailed knowledge of eligibility criteria and program scope was often lacking.

Awareness of the program was primarily shaped by informal and interpersonal channels. Dental clinics emerged as the most common source of information, followed by family and friends, while fewer participants reported learning about the benefit through government communications or media sources. As one participant explained, *“we visited [the] dental clinic… and at the reception we heard about the program”* (parent #13GUD).

Participants also highlighted inequities in awareness, noting that individuals with limited internet access or lower socioeconomic status may be less likely to receive information about the program. This raised concerns about whether this federal benefit was reaching those most in need.

### Theme 2: Inadequate and Inequitable Communication Strategies

Many participants perceived the federal government’s communication regarding the Interim CDB as insufficient, unclear, and poorly disseminated. While a small number of participants described the information as clear, the majority reported limited exposure to program information and uncertainty about how the benefit worked.

Communication was often described as passive, relying heavily on mailed letters or online platforms. Participants expressed concern that such approaches excluded individuals without reliable internet access, like those from rural and remote regions or those less engaged with formal communication channels. One participant noted, *“I haven’t seen any information on it… I’m not sure I would ever [have] heard about it”* (parent #3AR).

Additionally, participants reported that available information was often complex and difficult to interpret. Confusion around eligibility criteria, financial cut-offs, and application requirements further contributed to uncertainty and reduced engagement with the program.

### Theme 3: Barriers to Accessing the Interim CDB

Despite the availability of financial support, several barriers limited participants’ ability to access the benefit. A key barrier was the misunderstanding or misperception of eligibility criteria. Some participants believed they did not qualify due to factors such as income status or existing government supports, even when this was not accurate.

Administrative challenges were also frequently reported. Participants described the application process as complex, time-consuming, and difficult to navigate, even among those with higher levels of education. One participant reflected, *“it is not something easy even for someone who is knowledgeable… what about the majority?”* (parent #4DG).

Additional barriers included long wait times for telephone support, language challenges, and limited digital literacy. In some cases, participants missed application deadlines due to unclear timelines or a lack of awareness about program requirements, further limiting access.

### Theme 4: Dental Providers as Key Facilitators of Access

Dental professionals played a critical role in facilitating access to the Interim CDB. Most participants described dental clinic staff as knowledgeable, supportive, and instrumental in helping them understand and apply for the benefit.

Clinics served not only as points of care but also as key sources of information and guidance. Participants frequently credited dental providers and dental office staff with raising their awareness and assisting with navigation of the application process. As one participant stated, *“they gave me the information… and what to do with it”* (parent #21TA).

### Theme 5: Financial Relief and Improved Access to Care

The Interim CDB was widely perceived as beneficial in reducing the financial burden associated with dental care. Participants emphasized that the benefit enabled them to access services for their children that they might otherwise have delayed or foregone due to cost.

This was particularly evident among families with multiple children, where cumulative dental expenses can be substantial. One participant shared, *“it helped… [it] took the burden out of my pocket”* (parent #15JB).

### Theme 6: Gaps in Coverage and Ongoing Financial Strain

Despite its benefits, participants identified several limitations related to the scope and adequacy of the Interim CDB. While some participants felt the financial support was sufficient, others reported that it covered only basic services and did not cover the full cost of more complex treatments.

Perceptions of adequacy varied depending on individual circumstances and treatment needs. Some participants expressed uncertainty about whether the benefit was sufficient, noting that it “depends” on the child’s dental needs.

Participants also reported confusion about the benefit’s structure, including whether funding applied per child or per family and which services were covered. These uncertainties contributed to ongoing financial stress and challenges in planning care.

### Theme 7: Participant-Driven Recommendations for Program Improvement

Participants offered several recommendations to improve the effectiveness and reach of the Interim CDB. A key suggestion was to expand eligibility criteria, particularly by raising the age limit to include children up to 18 years old. Many participants felt that older children have greater dental needs and should not be excluded.

Another common recommendation was to increase the amount of financial support provided to better reflect the actual cost of dental care. Participants also emphasized the need for improved government communication strategies, including broader public awareness campaigns and the use of multiple dissemination channels.

Simplifying the application process and providing information in multiple languages were also highlighted as important steps to improve accessibility. Additionally, participants called for greater consideration of vulnerable populations, including those facing socioeconomic and structural barriers to care.

## Discussion

This study provides in-depth qualitative insights into parents’ experiences with the Canadian federal government’s Interim CDB in Manitoba, Canada. The findings highlight a complex interplay among awareness, communication, access, and financial impact, revealing both the program’s strengths and critical gaps that may influence its effectiveness and equity. Overall, while the Interim CDB was perceived as a valuable initiative that reduced financial barriers to care, important challenges related to awareness, accessibility, and adequacy of coverage remain.

A key finding of this study was the limited and uneven awareness of the Interim CDB among participants. Although many parents had some understanding of the program, knowledge was often partial and conveyed through informal channels such as dental clinics and social networks rather than through direct government communication. While the Government of Canada disseminated information through multiple avenues, including official websites, advertising campaigns, Canada Revenue Agency communications, and stakeholder engagement with dental associations and community partners (Government of Canada, 2022), our findings suggest that these strategies did not consistently translate into meaningful awareness at the community level. Instead, most participants reported learning about the program through dental offices or interpersonal networks. This finding is consistent with evidence from a survey of Manitoban parents, which similarly identified dental offices and family or friends as the most common sources of information and awareness regarding the Interim CDB (Baltus et al., 2026b).

These findings align with emerging evidence demonstrating that awareness and uptake of the CDB are not uniform across populations, with disparities observed among uninsured and socioeconomically disadvantaged groups (Goubran et al., 2024; Schroth et al., 2023). Similarly, Menon et al. (2026) reported that while six in ten parents had heard of the Interim CDB, only 22.4% felt well informed about the program. Regional differences were also evident, with higher levels of unawareness reported in Manitoba/Saskatchewan (37.8%) and Atlantic Canada (37.5%). Interestingly, although awareness was slightly higher among rural populations compared to urban populations, rural parents were less likely to apply for the benefit.

Taken together, these findings suggest that awareness alone is insufficient to ensure program uptake. The observed gap between awareness, understanding, and application likely reflects broader structural and informational barriers, including the complexity of program messaging, reliance on passive and digitally mediated communication strategies, and limited accessibility of information for populations with lower health literacy or reduced access to digital resources (Fitzpatrick, 2023). In this context, dental providers appear to play a critical intermediary role in translating policy into practice, bridging gaps between formal program communication and real-world access. This pattern is consistent with prior work demonstrating that parents often rely on dental professionals and familiar communication channels for oral health information. For example, in a previous study, a greater proportion of mothers reported receiving oral health information from dental providers, with preferred modes of communication including social media, television, and radio (Olatosi et al., 2021). Collectively, these findings underscore the need for more proactive, accessible, and community-engaged communication strategies to ensure equitable reach and effective implementation.

Closely related to this was the finding that communication strategies were perceived as inadequate and inequitable. Participants frequently reported limited visibility into the program and challenges understanding eligibility criteria and application requirements. Reliance on digital and passive communication methods, such as email or mailed notices, was viewed as insufficient, particularly for individuals with limited internet access or lower health literacy.

These findings highlight the importance of explicitly considering socially and structurally disadvantaged populations in the design and dissemination of public health programs, as these groups are often those most in need of care (Brondani et al., 2025).

Improving communication strategies will require a shift toward more inclusive and user-centered approaches. This includes developing culturally appropriate and linguistically accessible health information, tailoring messages to diverse audiences, and leveraging multiple communication platforms that align with community preferences. Effective communication should ensure that information is not only available, but also understandable and actionable across varying levels of health literacy (Fitzpatrick, 2023). Engaging target populations through trusted channels such as healthcare providers, community organizations, and widely used media platforms may further enhance reach and uptake (Menon et al., 2024). Ultimately, strengthening health communication practices will be essential to improving the effectiveness, equity, and impact of publicly funded dental programs.

Despite the availability of financial support, participants in this study reported several barriers to accessing the Interim CDB, including misconceptions about eligibility, complex application processes, and administrative challenges such as long wait times and unclear deadlines. Notably, some participants self-excluded based on incorrect assumptions about eligibility, highlighting the important role of perceived barriers in limiting program uptake. Several parents were unaware that they could apply for the Interim CDB if they held other government dental benefits for their children, such as the Non-Insured Health Benefits for Registered First Nations and Inuit Peoples or provincial social assistance. Similar challenges have been reported in studies examining both parent and provider perspectives, where administrative complexity and limited system navigation support were identified as key barriers to accessing the CDB (Baltus et al., 2026a; Menon et al., 2026). These findings suggest that beyond financial provision, program design and usability are critical determinants of access, underscoring the need for simplified application processes and enhanced support mechanisms.

An important and encouraging finding was the role of dental providers as key facilitators of access. Participants consistently identified dental clinic staff as trusted sources of information who played a central role in raising awareness and supporting navigation of the benefit. This finding is supported by evidence from a survey of oral health professionals in Manitoba, which found that dentists and dental receptionists were among the most influential in encouraging parents to apply for the Interim CDB (Baltus et al., 2026a). Together, these findings highlight the critical role of dental teams not only in delivering care but also in facilitating access to publicly funded programs. Leveraging dental clinics as points of engagement, education, and support may enhance uptake and reduce barriers, particularly among families already accessing care.

However, this also raises important considerations for reaching individuals who are not currently engaged with dental services and may remain excluded from such support pathways. Without complementary outreach strategies, reliance on dental settings alone may inadvertently reinforce existing inequities in access to care.

Consistent with the program’s intended purpose, the Interim CDB was widely perceived as providing meaningful financial relief and improving access to dental care. Participants described being able to seek care for their children that might otherwise have been delayed or unaffordable, particularly in families with multiple children. These findings align with emerging quantitative evidence indicating that the CDB has contributed to increased access to dental services among eligible populations (Goubran et al., 2024; Goubran et al., 2026; Schroth et al., 2023). However, the extent of this impact was moderated by concerns regarding the adequacy of coverage.

Indeed, gaps in coverage and ongoing financial strain emerged as a key theme. While some participants reported that the benefit was sufficient for basic care, others noted that it did not fully cover the cost of more complex treatments, resulting in continued out-of-pocket expenses. Variability in perceived adequacy reflects differences in treatment needs and underscores the limitations of fixed financial benefits in meeting diverse oral health needs. In addition, uncertainty regarding the structure and scope of the benefit contributed to confusion and challenges in care planning. These findings suggest that while the Interim CDB represents an important step forward, further refinement is needed to ensure more comprehensive, predictable, and needs-responsive coverage.

Participants also provided clear, actionable recommendations to improve the program, including expanding eligibility to older children, increasing financial support, improving communication strategies, and simplifying the application process. Notably, many participants emphasized the need for more inclusive and equitable approaches, including multilingual resources and targeted outreach to underserved populations. These recommendations are particularly relevant in the context of the ongoing implementation of the CDCP, which aims to expand access to dental care across Canada.

Taken together, these findings have important implications for policy and practice. As the CDCP continues to be rolled out, lessons learned from the Interim CDB can inform more effective and equitable implementation strategies. In particular, there is a need to prioritize clear, accessible, and culturally appropriate communication, streamline administrative processes, and ensure that financial supports align with the actual cost of care. Additionally, integrating oral health providers and community-based organizations into program delivery may enhance reach and uptake, particularly among populations facing structural barriers to care. Without such targeted and inclusive approaches, publicly funded dental programs may risk inadvertently perpetuating existing inequities rather than reducing them.

## Strengths and Limitations

This study provides rich, in-depth insights into parents’ experiences with a newly implemented national dental benefit, contributing important qualitative evidence to an emerging area of research. By capturing diverse perspectives, the findings offer valuable contextual understanding that complements existing quantitative analyses of program uptake and utilization. However, several limitations should be considered. The study was conducted in a single province, which may limit the transferability of findings to other regions with different population characteristics or service delivery contexts. Participants were primarily recruited through dental public health clinics, which may have led to the inclusion of individuals already engaged with oral health services and may not fully reflect the experiences of those facing the greatest barriers to care.

Additionally, as with all qualitative research, findings are context-specific and are not intended to be generalizable, but rather to provide depth of understanding and insight into participant experiences.

## Conclusion

The Interim CDB represents a significant step toward improving access to dental care for children in Canada. While the program has provided meaningful financial support and facilitated access to care, this study highlights important gaps in awareness, communication, accessibility, and adequacy of coverage. Addressing these challenges will be critical to ensuring the success of the CDCP and advancing equitable access to oral health care for all children.

## Supporting information

Supplemental File 1 - Questionnaire and Prompts

## Data Availability

All data produced in the present study are available upon reasonable request to the authors

## References

Baltus, T. H. L., Abdulrahman, A., Fux, S., Goubran, S., Mittermuller, B. A., Yerex, K., Menon, A., Monayao, A., Hai-Santiago, K., Singhal, S., & Schroth, R. J. (2026a). Oral Health Professionals’ Views on the Interim Canada Dental Benefit in Manitoba, Canada. J Public Health Dent. 10.1111/jphd.70047

Baltus, T. H. L., Youssef, C., Goubran, S., Mauli, G., Patel, D., Fux, S., Mittermuller, B.-A., DeMaré, D., Menon, A., Yerex, K., Hai-Santiago, K., Singhal, S., & Schroth, R. J. (2026b). Parents’ Awareness of, Views on and Experiences with the Interim Canada Dental Benefit. medRxiv, 2026.2002.2005.26345648. 10.64898/2026.02.05.26345648

Brondani, M. A., Jessani, A., Faria, E. S. A. L., & Ardenghi, D. M. (2025). Initial Findings from the Canadian Dental Care Plan: Policy in Action. J Can Dent Assoc, 91, 15.

Canada Revenue Agency. (2022). Canada Dental Benefit. Ottawa, ON: Government of Canada.

Fitzpatrick, P. J. (2023). Improving health literacy using the power of digital communications to achieve better health outcomes for patients and practitioners. Front Digit Health, 5, 1264780. 10.3389/fdgth.2023.1264780

Gondro, J. V., Emode, M., Ivancevic, D., Clarke, J., Ortlieb, K., & Farmer, J. (2025). Characteristics of cost-related avoidance of oral health services among people in Canada eligible for the Canadian Dental Care Plan. Health Rep, 36(8), 16–28. 10.25318/82-003-x202500800002-eng

Goubran, S., Cruz de Jesus, V., Menon, A., Olatosi, O. O., & Schroth, R. J. (2024). Uptake of the Interim Canada Dental Benefit: an investigation of data from the first 18 months of the program. Front Oral Health, 5, 1481423. 10.3389/froh.2024.1481423

Goubran, S., Youssef, C., Menon, A., Cruz de Jesus, V., Olatosi, O. O., Lee, V. H. K., & Schroth, R. J. A National Review of the Interim Canada Dental Benefit: Analyzing the Uptake Among Uninsured Children Under 12. Journal of Public Health Dentistry, n/a(n/a). 10.1111/jphd.70053

Goubran, S., Youssef, C., Menon, A., Cruz de Jesus, V., Olatosi, O. O., Lee, V. H. K., & Schroth, R. J. (2026). A National Review of the Interim Canada Dental Benefit: Analyzing the Uptake Among Uninsured Children Under 12. Journal of Public Health Dentistry, n/a(n/a). 10.1111/jphd.70053

Government of Canada. (2022). Question Period Note: Dental Care - Marketing and Advertising for the Interim Canada Dental Benefit.

Government of Canada. Government Bill (House of Commons). (2022). Government Bill (House of Commons) C-31—Royal Assent-An Act respecting Cost of Living Relief measures Related to Dental Care and Rental Housing.

Han, S. Y., Chang, C. L., Wang, Y. L., Wang, C. S., Lee, W. J., Vo, T. T. T., Chen, Y. L., Cheng, C. Y., & Lee, I. T. (2025). A Narrative Review on Advancing Pediatric Oral Health: Comprehensive Strategies for the Prevention and Management of Dental Challenges in Children. Children (Basel), 12(3). 10.3390/children12030286

Levy, B. B., Goodman, J., & Eskander, A. (2023). Oral healthcare disparities in Canada: filling in the gaps. Can J Public Health, 114(1), 139–145. 10.17269/s41997-022-00692-y

Menon, A., Cruz de Jesus, V., Virtanen, J. I., & Schroth, R. J. (2026). Parents’ Views on Access to Dental Care and the Interim Canada Dental Benefit. JDR Clin Trans Res, 11(2), 257–269. 10.1177/23800844251323169

Menon, A., Schroth, R. J., Hai-Santiago, K., Yerex, K., & Bertone, M. (2024). The Canadian dental care plan and the senior population. Front Oral Health, 5, 1385482. 10.3389/froh.2024.1385482

Olatosi, O. O., Mgbemere, O. J., Oyapero, A., Omotuyole, A. S., & Okolo, C. (2021). Awareness and preferred mode of getting information on first aid management of avulsed permanent teeth: survey of Nigerian mothers. Pesqui Bras Odontopediatria Clín Integr, 21:e0124

Schroth, R. J., Cruz de Jesus, V., Menon, A., Olatosi, O. O., Lee, V. H. K., Yerex, K., Hai-Santiago, K., & DeMare, D. (2023). An investigation of data from the first year of the interim Canada Dental Benefit for children <12 years of age. Front Oral Health, 4, 1328491. 10.3389/froh.2023.1328491

Schroth, R. J., Youssef, C., Cruz de Jesus, V., Olatosi, O. O., Lee, V. H. K., Goubran, S., Khan, E., & Menon, A. (2025). An investigation of the reach of the interim Canada dental benefit for children under 12 years of age [Original Research]. FRONTIERS IN ORAL HEALTH, Volume 6-2025. 10.3389/froh.2025.1611815

Tungare, S., & Paranjpe, A. G. (2025). Early Childhood Caries. In StatPearls.

