## Supplemental File 1 - Questionnaire and Prompts for "Bridging Policy and Practice: Parents’ and Caregivers’ Experiences with the Interim Canada Dental Benefit in Canada"

**Interim Canada Dental Benefit Evaluation**
**Parent/Caregiver Focus Group/Interview Guide**
**Qualitative Questions (open-ended)**

1. Can you tell me what do you know about the Interim Canada Dental Benefit?
   **Prompt**: how did you hear about the CDB? (CRA letter, TV/Radio, family member, friends, online, dental office, social media)

   **Prompt**: children under 12, child tax benefit, filed taxes in past two years, no private insurance,

2. How would you describe the information the public is receiving about the Interim Canada Dental Benefit from the Canadian government?

3. If you didn't apply for the Interim Canada Dental Benefit, can you tell me why?

4. If you were denied the Interim Canada Dental Benefit, can you tell us why?

5. How helpful/knowledgeable were the staff at the dental office/clinic you were taking your children to about the Interim Canada Dental Benefit?

6. What concerns about the Interim Canada Dental Benefit do you have?

7. Can you tell me how the Interim Canada Dental Benefit has helped you access oral health care for your child/children?

8. Do you feel that the amount you received was enough to cover the dental costs for your child?

9. What types of challenges did you encounter when applying for the interim Canada Dental Benefit?

10. What types of challenges did you encounter using the interim Canada Dental Benefit at a dental clinic?

11. What suggestions would you have to make the Interim Canada Dental Benefit better?
    **Prompt**: Would you recommend having the application information available in multiple languages to help address this potential barrier?
